# Characterising factors underlying praxis deficits in chronic left hemisphere stroke patients

**DOI:** 10.1101/2020.10.16.20213744

**Authors:** Elisabeth Rounis, Ajay Halai, Gloria Pizzamiglio, Matthew A. Lambon Ralph

## Abstract

Limb apraxia, a disorder of skilled action not consequent on primary motor or sensory deficits, has traditionally been defined according to errors patients make on neuropsychological tasks. Previous models of the disorder have failed to provide a unified account of patients’ deficits, due to heterogeneity in the patients and tasks used. In this study we implemented principal component analysis (PCA) to elucidate core factors of the disorder in a cohort of 41 unselected left hemisphere chronic stroke patients who were tested on a comprehensive and validated apraxia screen. Three principal components were identified: posture selection, semantic control and multi-demand sequencing. These were submitted to a lesion symptom mapping (VBCM) analysis in a subset of 24 patients, controlled for lesion volume, age and time post-stroke. Although the first component revealed no significant structural correlates, the second and third components were related to regions in the ‘ventro-dorsal’ and ‘ventral’ and ‘dorsal’ pathways, respectively. These results challenge the previously reported distinction between ideomotor and ideational deficits and highlight a significant role of common cognitive functions in the disorder, which include action selection, semantic retrieval, sequencing and response inhibition. Further research using this technique would help elucidate the cognitive processes underlying limb apraxia and their relationship with other cognitive disorders.

## 1. Introduction

Limb apraxia (thereafter referred to as ‘apraxia’) refers to a range of higher-order motor disorders resulting from acquired brain diseases that affect performance of skilled action not attributable to elemental sensory or motor impairments or to lack of comprehension (Heilman and Rothi, 1991, 2003). It is a common consequence of stroke, estimated to affect up to 50% of patients with left hemisphere lesions (Buxbaum et al. 2008, Zwinkels et al. 2004). In addition to stroke, apraxia is found in several other neurological conditions, with evidence to suggest that presence of apraxia is associated with an increased burden of disease (Donkervoort et al. 2006, Crutch et al. 2007, Bickerton et al. 2012). Patients with this disorder can be impaired in everyday tasks such as making a cup of tea. Apraxia may slow down motor rehabilitation in stroke patients with hemiparesis yet cause motor impairments in patients who seem to have recovered their hand function, with impairments affecting the ipsi-lesional, as well as the contra-lesional sides (Smania et al. 2000, Chestnut and Haaland 2008).

Despite its importance and a long history of reporting on the disorder, apraxia remains poorly understood. Original studies categorised apraxia on the basis of a two-system model of action organization: a conceptual and a production system (Liepmann 1908, Leiguarda and Marsden 2000). Classical work by Liepmann (1908, 1920) examined errors patients made in neuropsychological tasks. He attributed these errors either to deficits in conceptualizing the visual representations of ‘spatio-temporal plans’ of an action, which he termed ‘ideational’ (or ‘conceptual’) apraxia (Gerschwind and Damasio, 1985, Roy and Square 1985); or to deficits in implementing them, due to difficulty in retrieving or integrating them into a movement, which he termed ‘ideomotor’ apraxia (Leiguarda and Marsden 2000, Buxbaum and Randerath 2018). He hypothesized a role for extrastriate visual areas in representing the ‘idea of a movement’ (spatio-temporal plans), which required to be transferred from posterior to anterior areas of the brain for their implementation via the motor cortex. According to this, damage to the left parietal lobe was involved in ‘ideomotor’ deficits as it transforms movement ideas into ‘innervatory engrams’, which were implemented within sensory-motor areas bilaterally (Leiguarda and Marsden 2000, Goldenberg, 2009). He termed ‘limb-kinetic’ apraxia a further deficit in transforming the movement plans into an appropriate selection of muscle synergies to perform a skilled movement (Liepmann, 1920). Liepmann’s accounts of the disorder placed apraxia in the realm of ‘disconnection’ syndromes (Catani and Ffytche 2005).

Although Liepmann’s classification has remained in the literature, it has been widely debated due to inconsistencies in the terms used and the observation that both ideomotor and ideational deficits may co-occur in the same patients (Buxbaum 2001, Smania et al. 2000, Goldenberg 2013). More recent lesion-mapping studies have taken a different approach in investigating this disorder. This has been to base the neural deficits observed in apraxia on the dual stream model for visual organisation, which has identified a dedicated network of brain regions involved in skilled actions and object-use in humans and non-human primates (Mishkin et al. 1983, Goodale and Milner 1992, Leiguarda and Marsden 2000, Rizzolatti and Matelli 2003, Drapati and Sirigu 2006, Binkofski and Buxbaum 2013). According to this, the visual system is subdividied into a ‘dorsal’ stream representing visual control of actions and a ‘ventral’ stream, mediating object recognition (Goodale and Milner, 1992). The identification of parallel fronto-parietal pathways that represent specific properties of objects or the environment for sensorimotor integration and the generation of an appropriate movement has led to further subdivision of the dorsal pathway into a ‘ventro-dorsal’ fronto-parietal stream, comprising the inferior parietal lobule (representing intrinsic properties of objects such as their size or shape; Jeannerod, 1994) and ventral premotor cortex (mediating grasp selection; Fagg and Arbib, 1998). This interacts with a ‘dorso-dorsal’ stream, comprising superior parietal and dorsal premotor areas, implicated in on-line reaching and action selection (Rizzolatti and Matelli 2003, Cisek 2005). The degree of interaction between these fronto-parietal networks depend on task demands, the degree of online motor control and visual feedback required by an action (van Polanen and Davare, 2015).

Several lesion mapping studies report lesions within these networks relating to apraxia (Pazzaglia et al. 2008, Kalenine et al. 2010, Manuel et al. 2012, Hoeren et al. 2014, Buxbaum et al. 2014). However, even though this framework has been suitable in studies investigating detailed deficits in reaching and grasping tasks (Buxbaum et al. 2014), it does not fully account for findings in other aspects of apraxia testing, such as sequencing involving dorsal stream areas (Haaland and Harrington 1992), or pantomime deficits involving ventral stream areas (Finkel et al. 2018,

Pizzamiglio et al. 2019). In some tasks, both dorsal and ventral stream lesions appear to be involved in apraxia (Vry et al 2012, Buxbaum et al. 2014). There is increasing evidence that the ventral stream contributes important motor control functions. It plays a role in the representation of postures and object-hand interactions (Lingnau and Downing 2015, Bracci et al. 2013, 2018) and in action selection (Milner and Goodale, 2008; Weiller et al., 2009; Rijntjes et al., 2012). Recent DTI work demonstrating anatomical connections between the two streams, suggests greater integration between them than previously thought (Catani & Ffytsche 2005, Ramayya et al. 2010, Barbeau et al. 2020).

The behavioural tasks to evaluate conceptual and production deficits in limb apraxia have included pantomime of transitive (object-related) and intransitive (non-object related) movements as well as imitation of meaningful and meaningless gestures and postures for production deficits. Conceptual deficits, on the other hand, have been tested using action recognition tasks, variations of multi-step tasks (e.g., prepare a letter for posting), and tool selection or alternative tool selection tasks (Leiguarda and Marsden, 2000). Poeck (1986) encouraged the use of multi-step tasks in assessing apraxia, based on the observation that patients with apraxia exhibit several types of sequential errors. These have been attributed to frontal, parietal and basal ganglia lesions (Kimura and Archibald 1974, De Renzi et al 1983, Harrington and Haaland 1992, Halsband et al. 1993, Leiguarda and Marsden 2000).

Nevertheless, it is becoming increasingly apparent that tasks which aimed to separate deficits in apraxia involve common cognitive processes. These include, sequencing (noted above), response inhibition, perception of body in space (body schema – Goldenberg 2009), semantic representations (Hodges et al. 2000, Bozeat et al. 2002), semantic control (Corbett et al. 2009 and 2011, Watson and Buxbaum 2015), to name a few. Consequently, brain regions identified in deficits tested in one task may have more generic roles in praxis and in other cognitive processes. An example of this is the role of the parietal lobe, which is ubiquitous in very different cognitive functions (Humphreys and Lambon Ralph 2015), forming part of a ‘multi-demand’ system (Duncan 2010). Nevertheless, other brain areas are likely to represent functions that are task-specific and do not generalise. An example of this is the lateral occipito-temporal region, which has recently been identified to play a role in object-related and hand-posture representations (Bracci et al. 2013, 2018, Buxbaum et al. 2014).

Taken together, apraxia research has been limited by the fact that patients often do not conform either to previously defined ideational or ideomotor subcategories or to clear-cut neuro-anatomical correlates within the dual stream hypothesis (Binkofski and Buxbaum 2013). Limitations in the models used to describe the disorder, compounded by variability in the tasks used between studies have made it difficult to identify pathways to recovery.

Studies investigating recovery of other cognitive deficits after stroke, such as language, have faced similar challenges. Patients may often perform at ceiling (or floor) on some batteries of tasks, requiring multiple (possibly redundant or auto-correlative) tests of the cognitive function of interest for additional sensitivity and reliability (Butler et al. 2014). One approach of analysing the co-linearity and interactions within complex neuropsychological datasets is to use data-driven methods, such as principal component analysis (PCA). This approach, used previously in neuropsychology to explore subtypes of Alzheimer’s disease (Becker et al. 1988, Lambon Ralph et al. 2003), has more recently been applied in post-stroke aphasia, to identify reliable factors underlying deficits observed in patients, placing individual cases relative to each other in a multi-dimensional model (Butler et al. 2014, Mirman et al. 2015, Halai et al. 2017, Schumacher et al. 2019) and allowing comparisons of graded variations across one or more patient groups (Ingram et al. 2020).

In this study we utilised PCA in chronic left hemisphere stroke patients, specifically to identify the underlying latent structure of apraxia and to map the neural correlates of the emergent components. This was achieved by testing patients using a detailed apraxia screen, derived from the Birmingham Cognitive Screen, which has been validated in stroke patients (Humphreys et al. 2012, Bickerton et al. 2012). We hypothesised one of two patterns of results. Either we might identify a distinction based on traditional ‘ideational’ or ‘ideomotor’ models of the disease, with tasks of gesture recognition, complex figure copy and multi object use falling in the former category and tasks of gesture production, single object use and response inhibition, in the latter. Alternatively, we hypothesised that the PCA might distinguish between ‘domain general’ action deficits, involving action selection retrieval and/or sequencing and ‘domain specific’ deficits relating to motor representations of familiar gestures and/or object use in which ideational and ideomotor deficits co-occur. The brain regions subserved by the latter would predict distributed areas within dorsal and ventral stream subdivisions (Pazzaglia et al. 2008, Kalenine et al. 2010, Hoeren et al. 2014, Buxbaum et al. 2014, Dressing et al. 2018, Wong et al. 2019).

## 2. Methods

### 2.1 Participants

Forty-six unselected left hemisphere chronic stroke patients (first ever ischaemic or haemorrhagic) were recruited in the study, forty-one of which were included in the analyses (age range 25-79y.o. mean=58y.o.; M=29; F=12). Two patients were excluded because they were left-handed, and three because they had bilateral lesions. All remaining patients (N=41) were pre-morbidly right-handed, native English speakers with normal or corrected-to-normal vision. Their mean no of years of education was 13.3 yrs (range 10-20years) and mean time post-stroke was 28 months (range 12-62months). Exclusion criteria for the study were previous strokes, right hemisphere lesion, left-handedness, significant cognitive impairment precluding the ability to understand and provide written informed consent, or any other neurological or psychiatric condition.

Full written consent according to the declaration of Helsinki was obtained from all participants. The study was approved by the Health Research Authority, South Central – Berkshire Ethics committee. The study procedures or analyses were not pre-registered prior to the research. Participants attended the Cognitive Neuropsychology Centre, at the Department of Psychology (University of Oxford), for a detailed neuropsychological testing session. A subset of twenty-four patients who had no contraindications for MR imaging attended a second session at the Oxford Centre for Magnetic Resonance Imaging at the University of Oxford for structural brain imaging.

MRI data from a healthy age and education matched control group (10 females, 12 males), acquired in previous studies (Butler et al. 2014, Halai et al. 2020), was used as a reference to identify lesion/abnormal tissue for each patient (Seghier et al. 2008).

### 2.2 Neuropsychological assessments

This study used the battery of praxis tasks for stroke developed as part of the Birmingham Cognitive Screen (Humphreys et al. 2012). This comprised gesture recognition and gesture pantomime, single object-use as well as meaningless gesture imitation. Additional tasks used from this screening tool included the multi-object use and complex figure copy tasks. We also tested for response inhibition using a go-no go task (Verbuggen and Logan, 2008). These tasks are described in greater detail, below.

#### i Multi-step Object U1se Task

In this task, patients were provided with the instruction to ‘make a torch work’. The task instruction was also provided with a photograph to ‘light the torch’. They were presented with target objects (namely two batteries and a torch) along with distractor objects (a glue stick, a screwdriver, and matches). The task required they place the batteries provided in the torch and light it. In patients with unilateral weakness, the examiner would provide help, for example, stabilizing the torch barrel on the patient’s request or when patients showed signs of initiating the appropriate action. The task was scored out of 12. The first eight points were administered for the correct completion of every step of the task (unscrewing the torch barrel with no prompts, filling the barrel, inserting the battery from the cylindrical opening, inserting two batteries, closing the barrel after the batteries were inserted, ensuring the top was inserted in the correct orientation, switching the torch on after closing the barrel, achieving the goal of lighting the torch). The latter four points corresponded to errors during the task, in which a score of 1 was given for no error and a score of 0 was given if there was an error (with no negative scores). The type of errors included: 1) the number of attempts to achieve the task (1, for up to 2 attempts, 0 if there were more than 2 attempts), 2) use of irrelevant objects (scored 1 if no irrelevant objects were used, 0 if the matches, screwdriver or the glue stick were used), 3) irrelevant actions with the target object (scored 1 if no irrelevant actions were observed, 0 if there were), 4) perseveration (scored 1 if perseveration was absent, 0 if it was present).

#### ii Response Inhibition measured using the Go/NoGo task

In this task, patients were presented with a total of 90 trials, with frequent ‘Go’ trials (60) and infrequent ‘NoGo’ trials (30). In each trial the patient was asked to focus on a central white fixation cross of a black PC monitor screen located 30 cm from the participant, for 1200ms. A green square was presented for 300ms in the centre of the screen, which was either followed by nothing (‘Go’ trial) or by a red circle (‘NoGo’ trial) presented for 1000ms. Participants were asked to press a response key as quickly as possible following the green square if it was not followed by a red circle (a ‘Go’ trial). However, if a red circle appeared after the green square, then participants had to withhold their response (‘No-Go’ trial). There were 3 inter-stimulus time intervals in NoGo trials between presentation of the green square and red circles of 150ms, 200ms or 250ms. There was a 2000ms inter-trial interval, from disappearance of target and the next trial. Errors of commission were noted when participants pressed the response key in a ‘No Go’ trial. These were noted to be more frequent at the 250ms time window, which is the error reported in this study. Errors of omission were noted when participants did not press the response key after a ‘Go’ trial.

#### iii. Gesture Recognition

In the Gesture Recognition Task, the examiner produced six actions, which patients had to recognise: three transitive (using a cup, using a key, and using a lighter) and three intransitive (come over, good, and goodbye) actions. The examiner showed each gesture while the patients had to select the action being performed from a multiple-choice list, which included four alternative responses for each action, in writing. The four alternatives for each action corresponded to: 1) the correct action (e.g., using a lighter); 2) a semantically related action (using a match); 3) a visually related action (using a gun); and 4) an unrelated action (using a torch). The patients were allowed a maximum of 15 seconds per item to respond by pointing to their chosen statement and they were given 1 point for each correct response. The total correct score (maximum=6) was used in the analyses. The data from both transitive and intransitive gestures in these tasks were entered together as a composite measure; hence, this study is not reporting differences between the two.

#### iv. Gesture Production

The Gesture Production task involved pantomime of a total of six gestures (three transitive and three intransitive) to verbal command. The test included body centered (salute and using a glass), non-body-centered (stop and using a salt cellar), repetitive (hitch-hiking and using a hammer), and non-repetitive (stop and using a glass) actions. All actions can be carried out as a single step sequence. Patients were allowed a maximum of 15 seconds per item to respond and were asked to execute the action once. Two points were given for a correct and accurate gesture; 1 point for a recognizable but inaccurate gesture (e.g., including spatial and/or movement errors); 0 points were given for either no response after 15 seconds, an unrecognizable response or perseveration from previous gestures. The total correct score (maximum=12) was used in the analyses.

#### v. Meaningless Gesture Imitation

The patients were asked to copy four meaningless hand gestures and six meaningless finger gestures presented by the examiner. Two points were given for a gesture that was correctly and precisely imitated after the 1^st^ presentation; 1 points if the gesture was correct and precise after the 2^nd^ presentation; 0 point if patients made no response or incorrectly imitated the gesture after the 2^nd^ presentation (e.g., incorrect spatial relationship between hand and head, or incorrect finger/hand position), or showed perseveration from previous item(s) after the 2^nd^ presentation. The total correct score (maximum=20) was used in the analyses.

#### vi. Single Object Use

In the Single Object Use task patients were presented with one of six objects individually, one at a time (torch, straw, comb, nail clipper, screwdriver and matches). They were asked to demonstrate use of each of these with the object at hand. The patients were allowed a maximum of 15 seconds per item to respond. Two points were given for a correct and accurate gesture; 1 point for a recognizable but inaccurate gesture (e.g., including spatial and/or movement errors); 0 points were given for either no response after 15 seconds, an unrecognizable response or perseveration from previous gestures. The total correct score (maximum=12) was used in the analyses.

Gestures for all six tasks mentioned above were videotaped and later coded as correct or incorrect according to the scoring system detailed for each task. Two independent coders (ER, GP) scored the videos for each participant and each task. The final score for each task consisted of the average between the two scores. The inter-coder reliability defined by Cohen’s Kappa was ‘moderate’0.77 (McHugh, 2012). It is noteworthy that patients included in this study were unselected such that coders were blind as to whether they had praxis deficits or not.

#### vii. The complex figure copy test

Patients copied a complex figure, from the BCoS (Humphreys et al. 2012), as accurately as possible (Chen et al. 2016). The BCoS complex figure task is very similar to the Rey-Osterrieth figure copy test (Rey, 1941, Osterrieth, 1944). The figure contains a middle structure, as well as structures to the left and the right, which combine a total of 16 features to be copied. Each feature is scored on three criteria: presence, shape and placement (noting the middle square consists only of the former 2 criteria). The total score corresponds to the sum of features that have been accurately reproduced, with a maximum score of 47.

### 2.3 Principal Component Analysis

The participants’ scores on all the tasks mentioned above were entered into a PCA with varimax rotation (SPSS 25.0). Forty-one patients were included with the following 8 variables: (i) multi-object use, (ii) go-nogo commission errors at 250ms, (iii) go-no go omission errors, (iv) gesture recognition, (v) gesture production, (vi) complex figure copy, (vii) single object use and (viii) meaningless gesture imitation, leading to a ratio of subject to variable of 5.1 (Barrett and Kline, 1981). Factors with an eigenvalue exceeding 1.0 were extracted and then rotated. Following orthogonal rotation, the loadings of each task enabled interpretation of cognitive or motor control processes underlying each factor. Individual patients’ scores on each extracted factor (using regression) were then used as behavioural covariates in the neuroimaging analysis.

### 2.3 Neuroimaging data acquisition

The patient scanning was undertaken on a Siemens 3T Trio MRI scanner at the University of Oxford Centre for Clinical Magnetic Resonance Research (OCMR), in a standard 12-channel head coil. High resolution structural T1-weighted MR images were acquired using the MP-RAGE sequence (repetition time, 2040ms; echo time 4.7ms; field of view 174×192mm^2^; 192 slices; voxel size, 1×1×1mm^3^, flip angle = 8°), the total scan acquisition time was 556 seconds. The imaging protocol included a Fluid-Attenuated Inversion Recovery (FLAIR) scan (TR: 9s, TE: 90ms, FOV 220 x 220mm, axial plane; slice thickness: 3mm, 47 slices).

The MRI scanning acquisition parameters for the healthy control group used for the imaging analyses are identical to that reported in Butler et al. (2014) and Halai et al. (2020).

### 2.5 Neuroimaging data analysis

Structural MRI scans were preprocessed using Statistical Parametric Mapping software (SPM12 Wellcome Trust Centre for Neuroimaging: http://www.fil.ion.ucl.ac.uk/spm/), running on Matlab 2017a (https://www.mathworks.com/). The images were normalized into standard Montreal Neurological Institute (MNI) space using a modified unified segmentation-normalisation procedure for focal lesioned brains (Seghier et al. 2008). For the manual lesion delineations, one author (GP) traced the lesions manually on patients’ individual T1 image in native space, while consulting the FLAIR coregistered sequence, using MRIcron (https://www.nitrc.org/projects/mricron). The lesions were identified on a slice by slice basis and were checked by a trained neurologist (ER) once completed. Binary masks were made from the lesions using MRIcron (Rorden et al. 2009). These were co-registered to T1 native space and the lesion mask was used during the segmentation-normalisation procedure as a cost-function mask (Brett et al. 2001). Data from all participants with stroke and healthy age matched controls were entered into the segmentation-normalisation. Images were smoothed with an 8 mm full-width at half-maximum (FWHM) Gaussian kernel and submitted to Seghier’s et al. (2008) automated routine’s lesion identification and definition modules using default settings except from the lesion definition ‘U-threshold’, which was set to 0.5 as in previous studies (Butler et al. 2014, Halai et al. 2017, Schumacher et al. 2019). The manual lesions in native space were normalised into MNI space using the deformation fields obtained during segmentation-normalisation and summed across all subjects with brain imaging to create a lesion overlap map (Figure 1).

**Figure 1:**
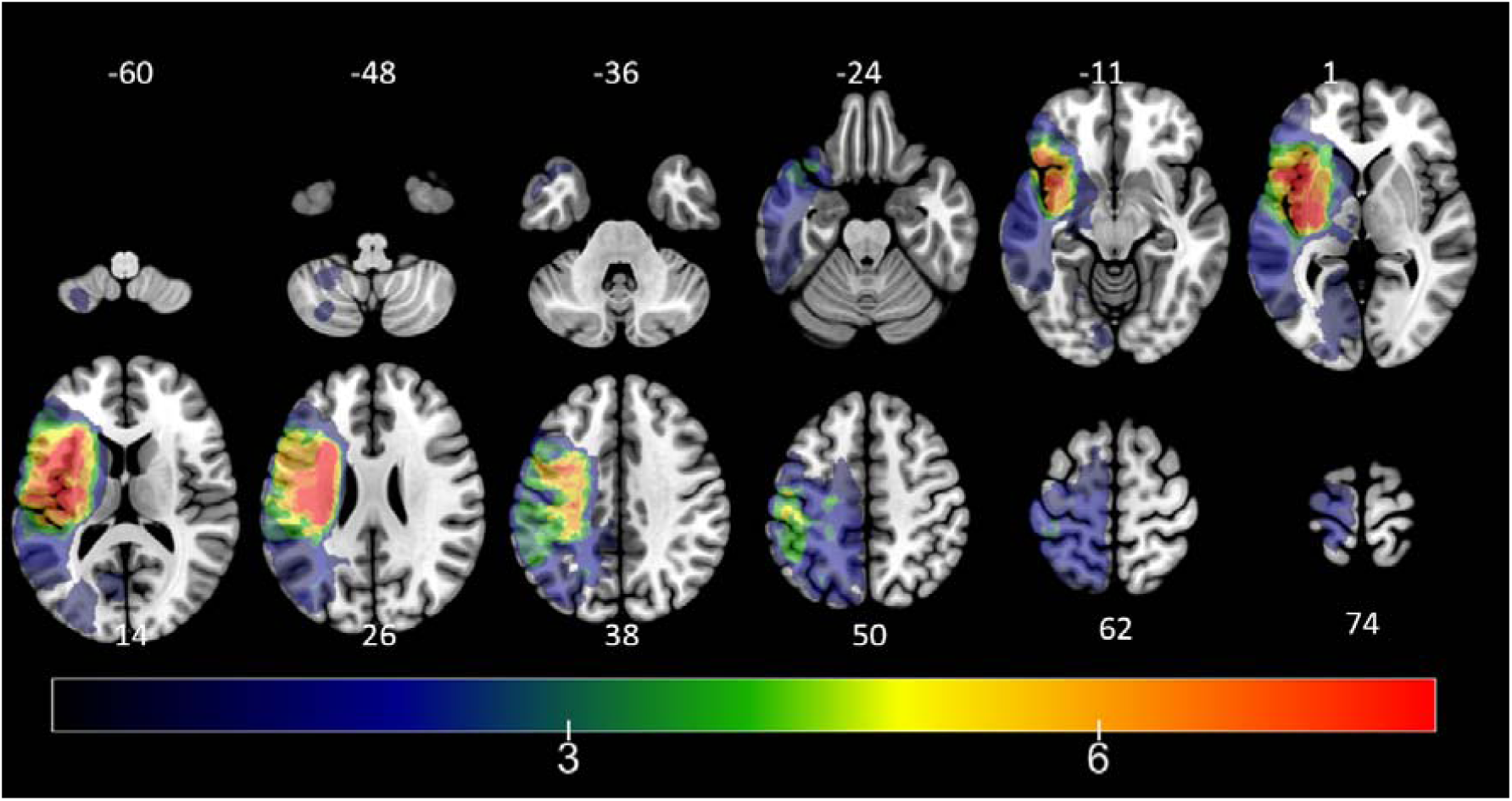
Lesion overlap map for 24 patients with left hemisphere stroke: The colour bar indicates the number of patients that had a lesion in each voxel [1-8]. The number adjacent to each brain slice is the Z (axial) coordinate in MNI space. (MRICroGL https://www.mccauslandcenter.sc.edu/mricrogl)

We took the smoothed T1-weighted images in MNI space (containing continuous signal intensity values across the whole brain) to determine the brain regions where ‘lesions’ (indicated by reduced signal intensity) correlated with PCA factor scores using a voxel-based correlational methodology (Tyler et al. 2005), a variant of voxel-lesion symptom mapping (VLSM: Bates et al. 2003), in which both brain and behavioural measures are continuous variables (conducted in SPM12). The participants’ component scores from the PCA were entered simultaneously into a VBCM analysis, with additional covariates controlling for age, time post-stroke, lesion volume (using manually delineated lesions) and mean signal in the right hemisphere. Statistical significance was determined by applying a voxel-level threshold of p < 0.001 and family-wise error corrected (FWEc) cluster-level threshold of p < 0.05.

The cortical anatomy of significant regions was identified based on the multi-modal parcellation (MMP) of human cerebral cortex provided by the Human Connectome Project (HCP) (Andreas, 2016; Glasser et al., 2016). The white matter tracts of significant clusters was based on The Atlas of Human Brain Connections (Rojkova et al, 2016). Figure 3 was created using the ‘ch2better’ template in MRIcron (www.nitrc.org).

**Figure 2:**
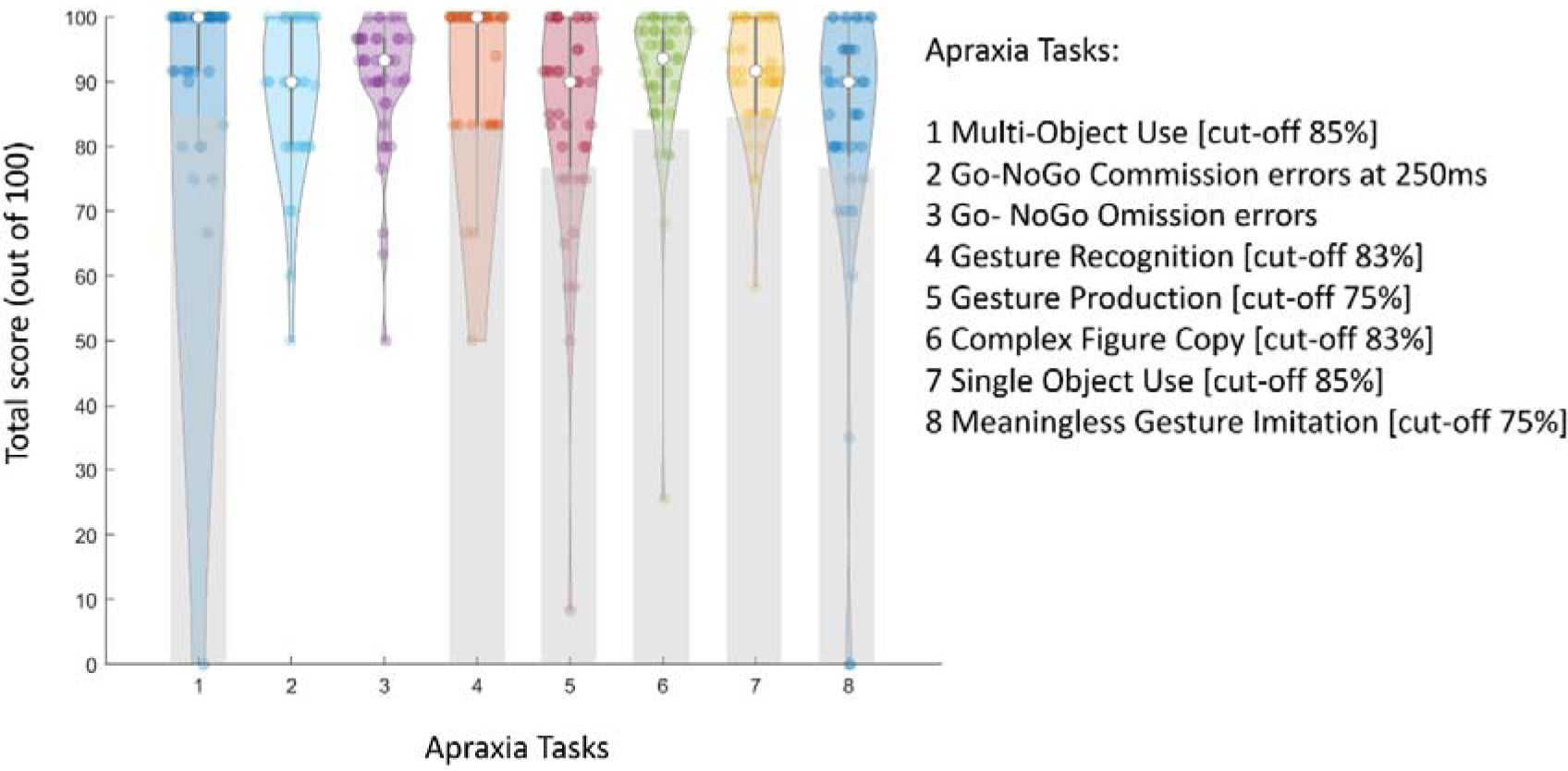
Patient performance in all apraxia tasks. Violin plots showing patient performance in each task included in the rotated PCA (note cut-off scores are highlighted in grey, from Humphreys et al. (2012). Each coloured dot represents a score from a subject and the white dot represent the median. Most patients performed near ceiling in tasks of multi-object use, and gesture recognition.

**Figure 3:**
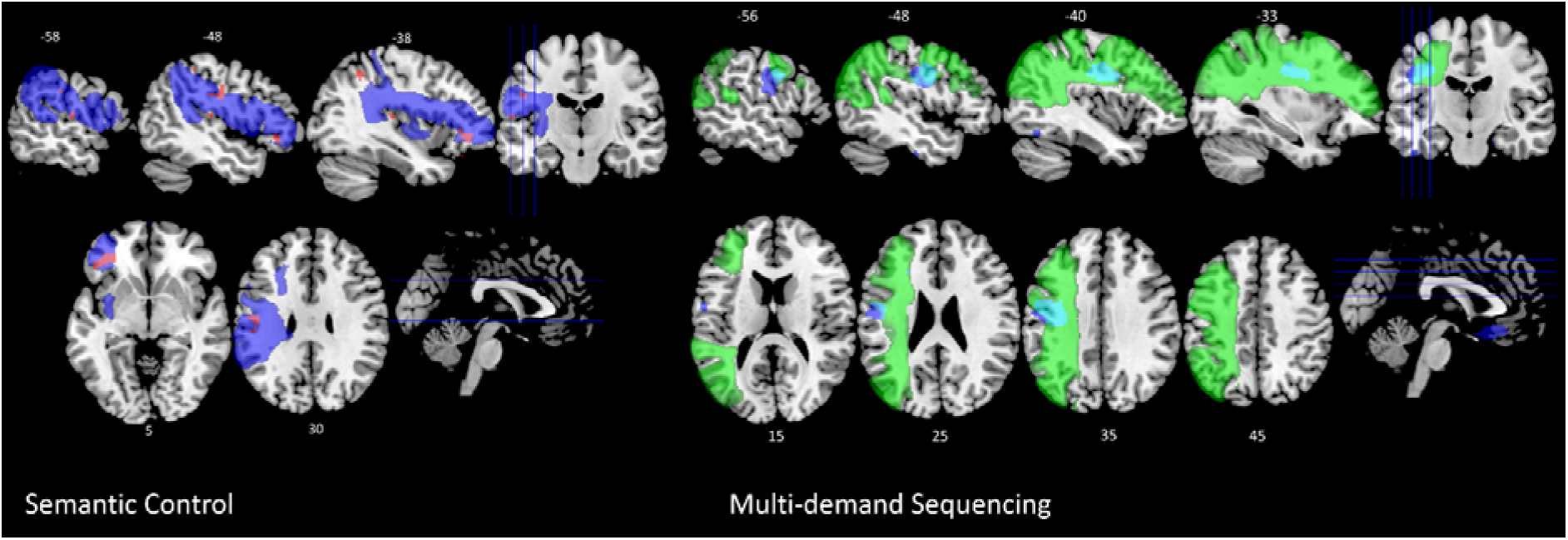
Significant lesion clusters for semantic control (left, red) and multi-demand sequencing (right, blue) VBCM in which age, time post-stroke, lesion volume and mean right hemisphere signal have been included as covariates. The figure on the left also shows the third branch of the Superior Longitudinal Fasciculus (blue) and overlap with significant clusters (pink). The figure on the right also shows the second branch of the Superior Longitudinal Fasciculus (green) and overlap with significant clusters (cyan). T-maps are thresholded at p<0.001, cluster corrected at FWE of p<0.05. The top row shows sagittal and bottom row axial views.

## 3. Results

### 3.1 Neuropsychological and lesion profiles

The group level scores for each behavioural task were as follows: multi-object use task (Mean=92.2, StDev=17.2, N=7/41 patients scored below normal cut-off), Go-NoGo commission (Mean=88.7, StDev=11.9), Go-NoGo omission (Mean=90, StDev=10.6), gesture recognition (Mean=93.3, StDev=11.7, N=9/41 patients scored below normal cut-off), gesture production (Mean=84.2, StDev=17.9, N=14/41 below cut-off), complex figure copy (Mean=90.7, StDev=13, N=6/41 scored below cut-off), single object use (Mean=90.9, StDev=8.9, N=10/41 scored below normal cut-off), meaningless gesture imitation (Mean=81.7, StDev=22.9, N=16/41 scored below cut-off). The individual scores on each task are shown in Supplementary Table 1.

Figure 1 shows a lesion overlap of the subset of 24 stroke patients with brain imaging, primarily affecting the MCA vascular territory within the left hemisphere (Phan et al. 2005). The maximum number of participants who had a lesion at any one site was 8 (located at x=-28, y=-9 z=1, within the left posterior putamen).

### 3.2 Identifying principal component factors underlying limb apraxia

The 8 variables entered into a varimax rotated PCA yielded three orthogonal components, accounting for 69% of the variance in patients’ performance [Kaiser-Meyer-Olkin (KMO) = 0.68]. Based on the factor loadings on each component (Table 1), the first component (accounting for 26.6% of the variance) was interpreted as ‘posture selection’. This component contained tests of response inhibition (the two measures of go-no go tasks) and meaningless gesture imitation. The second component, comprising the tasks of gesture recognition, gesture production and single object use was interpreted as ‘semantic control’ and accounted for 20.57% of variance. The third component was interpreted as ‘multi-demand sequencing’ and accounted for 12.51%. It included the multi-object use and complex figure copy tasks.

**Table 1:** Factor loadings for PCA of apraxia measures (Factor loadings exceeding 0.5 are marked in bold)

|  | FACTOR 1<br>Posture Selection | FACTOR 2<br>Semantic<br>Control | FACTOR 3<br>Multi-demand<br>Sequencing |
| --- | --- | --- | --- |
| Meaningless Gesture Imitation | **0.66** | 0.00 | 0.12 |
| Go-NoGo Commissions | **0.90** | 0.07 | -0.14 |
| Go-NoGo Omissions | **0.83** | 0.30 | -0.00 |
| Gesture Recognition | -0.08 | **0.91** | 0.01 |
| Gesture Production | 0.31 | **0.78** | 0.20 |
| Single Object Use | 0.35 | **0.61** | 0.43 |
| Figure Copy | -0.03 | 0.23 | **0.72** |
| Multi-Object Use | 0.01 | 0.01 | **0.83** |

### 3.3. Identifying lesion sites for action selection, action semantics and action sequencing

The VBCM analysis for each independent factor was carried out on the subset of 24 patients who underwent brain imaging. Significant clusters were identified for semantic control and multi-demand sequencing (Figure 3), with peak MNI coordinates shown in Table 4. The clusters identified areas where MR signal covaried with a given factor score, after accounting for lesion size, mean right hemisphere signal, age and time post-stroke. We did not identify any significant clusters for the ‘posture selection’ factor, which comprised the Go-No-Go and Meaningless Gesture Imitation tasks. The semantic control factor was uniquely correlated with voxels within part of the previously reported ‘ventro-dorsal’ stream (Binkofski and Buxbaum, 2013): 1) the ventral premotor area (Brodmann areas 45 – 47), within inferior frontal gyrus, adjacent to the left uncinate fasciculus and 2) the primary motor area, and adjacent parietal opercular, inferior parietal and supramarginal gyrus areas, abutting the third branch of the superior longitudinal fascicle (SLF III) (Figure 3, Rojkova et al. 2016). The multi-demand sequencing factor was associated with the white matter underlying the left sensorimotor cortex, involving the 2^nd^ branch of the left superior longitudinal fasciculus SLF II (Figure 3) and (bilateral, but predominantly-right) posterior orbitofrontal cortex (pOFC), adjacent to the right anterior cingulum bundle (Rojkova et al. 2016).

**Table 2:**
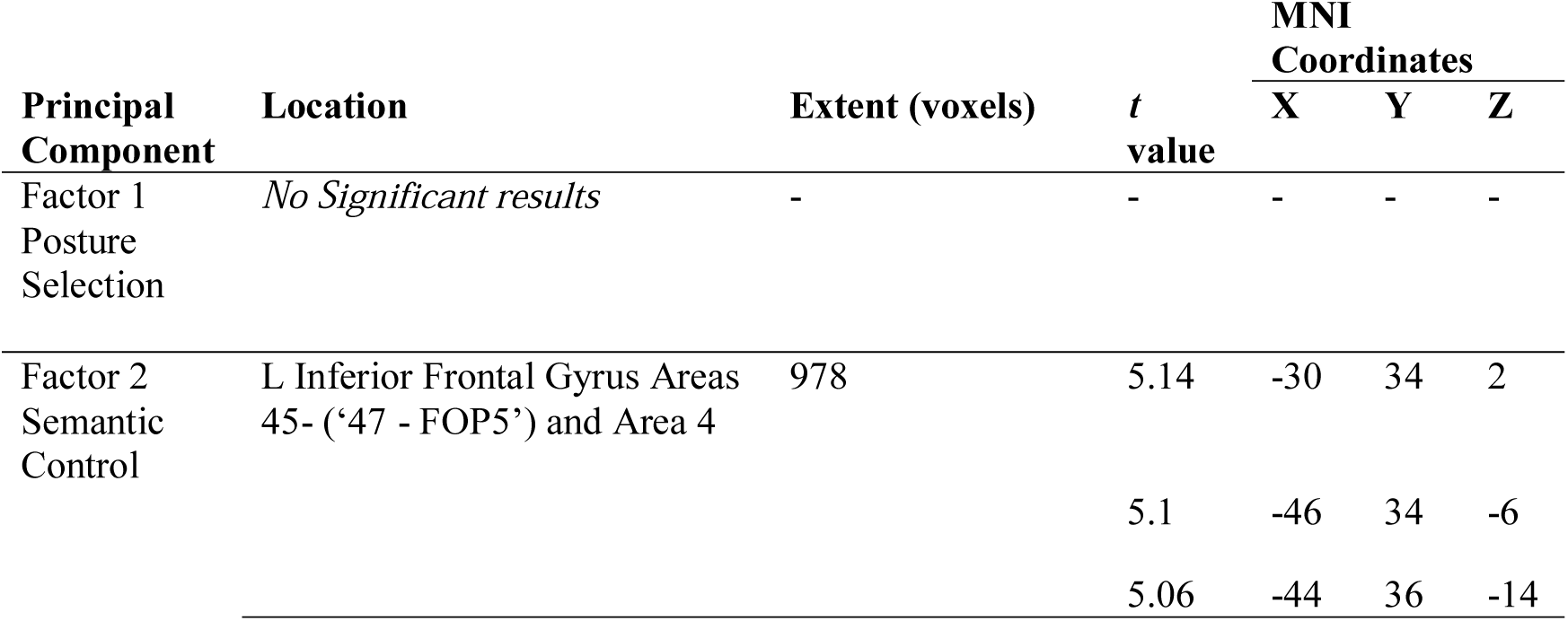
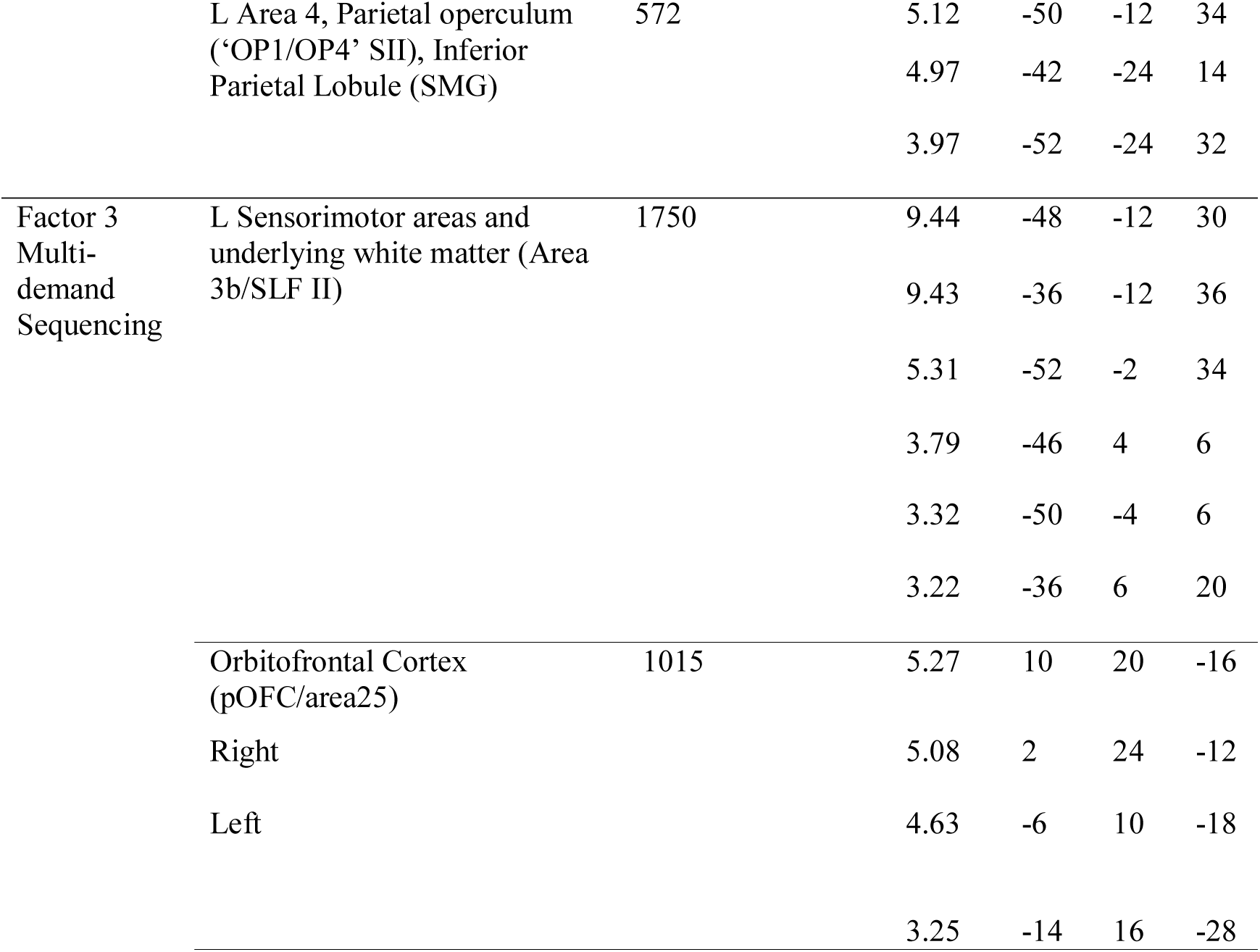
Details of significant VBCM results mapping principal components to brain damage. Peaks are reported in MNI space with anatomical labels from the MMP atlas and Catani atlas of human brain connections (de Schotten et al. 2011). Results were thresholded using p < 0.001 voxel-height and p < 0.05 FWE cluster-based correction.

## 4. Discussion

We report results on the use of PCA to study apraxia in an unselected cohort of chronic left hemisphere stroke patients. PCA was implemented on patients’ scores in six praxis tasks from the Birmingham Cognitive Screen (BCoS) (Humphreys et al. 2012, Bickerton et al., 2012) and two scores measuring motor response inhibition in a Go-NoGo task. In a subset of patients who had undergone MR imaging, we also conducted VBCM analyses in which brain damage was correlated with underlying factors of apraxia deficits, corrected for lesion volume, age and time post-stroke. The PCA identified three principle factors: 1) posture selection, 2) semantic control and 3) multi-demand sequencing. The latter two factors significantly mapped to neural damage falling within networks involving both the SLF II and SLF III, as well as the left uncinate and the anterior cingulum pathways respectively (Rojkova et al. 2016, Barbeau et al. 2020). A previous study used parallel analysis to distinguish apraxia and spatial inattention in left hemisphere stroke patients (Timpert et al. 2015). Although this preliminary technique was less robust in separating some of the components, such as imitation, and the VLSM results did not control for lesion volume, they were able to identify separable apraxia tasks localising within the left SLF, as in our study. These results corroborate original accounts of disconnection in this disorder (Liepmann 1908, 1920, Gazzaniga et al. 1967, Watson et al. 1986, Heilman and Watson 2008).

We discuss the rationale for using PCA in apraxia, how the results of this study relate to traditional tasks used in this disorder and the relationship of our imaging results with recent lesion symptom mapping literature of apraxia. We conclude with implications of using this technique to direct for future research in the field.

### The role of tasks used in apraxia models

As with other cognitive deficits, research in limb apraxia has been limited by several sources of variability that have made it difficult to provide a unified account of the disorder. There has been heterogeneity in the tasks used, the models accounting for limb apraxia and in the patient cohorts studied (in terms of their lesion characteristics, with selection of predominantly left MCA stroke patients in most studies, as well as the time they were studied since stroke (acute (Hoeren et al. 2014) vs chronic (Buxbaum et al. 2014, Wong et al. 2019)).

Nevertheless, the most important concern has been the use of different tasks across studies (Buxbaum and Randerath 2018). There are a few recognized screening assessments for apraxia which include a comprehensive number of tasks traditionally used in the disorder (‘BDAE’, Goodglass et al. 2000; ‘TULIA’, Vanbellingen et al. 2011; ‘BCOS’, Humphreys et al. 2012). It is noteworthy that most apraxia tasks, even ones considered as reflecting ‘direct’ pathways to motor execution, such as meaningless gesture imitation (Rothi et al. 1991), often comprise multiple cognitive processes. For example, meaningless gesture imitation requires ‘domain general’ executive processes such as attention, short term memory, task switching as well as domain specific ones, including 1) action observation (Allison et al. 2000, Saygin 2007), 2) transformation of a gesture into one’s own body schema (Wolpert and Haggard 2005), 3) organising movement according to a ‘hierarchy of goals’ (Bekkering et al. 2005), 4) selecting the correct finger or hand configurations which may be competing for action execution with others (requiring one to inhibiting unwanted actions) (Romaniuk, 2011), and 5) online self-monitoring, i.e. recognizing that the executed action matches what was originally shown (Blakemore et al. 2000). Moreover, there is recent evidence that meaningless imitation may use action recognition pathways (Pizzamiglio et al. 2019). A study in stroke patients, with and without apraxia, showed improved imitation of ‘meaningless’ gestures if they were perceived to be more familiar (Achilles et al. 2016).

In this study, we chose to address these issues by utilising PCA to uncover latent factors underlying apraxia by sampling a range of tasks. This has the benefit of capturing shared variance across a wide variety of tasks and distilling the core underlying features or processes of this disorder. This methodology has been applied, for example, to stroke aphasia and proved effective in identifying core components of language (phonology, semantics, speech output abilities) (Butler et al., 2014; Halai, et al., 2017) and executive function (shift-update, inhibit-generate, and speed of processing) (Schumacher et al. 2019).

The rotated PCA revealed three significant factors, confirming the utility of this approach in studying the disorder. The first, ‘posture selection’ factor included meaningless gesture imitation and Go-No Go tasks. This result suggests that selecting a particular finger or posture configuration in meaningless gesture imitation requires the inhibition of unwanted, possibly mutually competing, hand or finger gestures (Romaniuk 2011). Hence the inclusion of a Go-No Go task was able to distinguish this factor from others. Action selection deficits have been observed in several studies investigating apraxia in schizophrenia (Romaniuk 2011, Walther et al. 2013, 2020, Rounis and Humphreys 2015). In another psychiatric condition with similar response inhibition deficits, namely Obsessive-Compulsive Disorder, patients are also impaired in meaningless gesture imitation (Rounis et al. 2016). The second factor comprised tasks that have traditionally been attributed to ideomotor (gesture production, single object use) and ideational (gesture recognition) deficits (Leiguarda and Marsden 2000). It is interesting that as has been observed in previous studies, both were identified in the same factor (Buxbaum and Randerath, 2018). This factor was labelled ‘semantic control’, as it parallels semantic control deficits observed in the language domain where selecting or being cued with the appropriate context is important (Jefferies and Lambon Ralph 2006, Corbett et al 2009, Corbett et al. 2011). The behavioural and neuroanatomical underpinnings of these would involve cognitive processes that include object recognition (Mahon et al. 2007), action recognition (Pazzaglia et al. 2008, Urgesi et al. 2014) and the ability to combine them (Negri et al.2007), perhaps supported by executive/semantic related processes (Corbett et al. 2009 and 2011).

Finally, the third ‘multi-demand sequencing’ factor included complex figure copy and multi-object use tasks. These tasks have common underlying processes of requiring patients to hold multiple items in working memory and ordering them in sequence to complete a goal. It parallels the Naturalistic Action Task (Schwartz et al. 2010), which also involves several cognitive processes of executive function including sustained attention, working memory and sequencing in a ‘hierarchy of goals’ framework (Hamilton and Grafton, 2007). Several authors have argued for a ubiquitous role of sequencing deficits in apraxia (Poeck 1986, Harrington and Haaland, 1992). Patients with semantic control deficits have also been shown to have significant deficits in tasks similar to those observed in our patient cohort, which included fragmented action sequences, failure to complete subtasks or perseveration (Jefferies and Lambon Ralph 2006, Corbett et al. 2009, Noonan et al. 2013). Taken together the deficits observed in the second factor seem to represent an impairment in semantic control of actions and tool gestures (Buxbaum and Saffran 2002), whereas deficits relating to the third factor appeared to be related to domain general multi-demand impairments (Duncan et al 2010). This is corroborated by findings in the VBCM analysis, discussed below.

### The neural correlates of deficits underlying limb apraxia

Previous literature on the neural correlates of limb apraxia report a significant role for the ‘ventro-dorsal’ visuomotor stream in object use and action pantomime (Vry et al. 2012, Binkofski and Buxbaum 2013, Hoeren et al. 2014, Buxbaum et al. 2014, Dressing et al. 2019). This network has been identified both in humans and non-human primates as reported in the Introduction (Rizzolatti and Mattelli 2003, Drapati and Sirigu 2006). However, other apraxia tasks, such as meaningless gesture imitation (Buxbaum et al. 2014, Wong et al. 2019), naturalistic actions and multi-object use tend to be attributed to more distributed pathways (Tessari and Rumiati 2004, Hartmann et al. 2005, Goldenberg and Karnath 2006).

The ‘semantic control’ factor identified in this study’s PCA correlated to structural deficits in areas within left SLF III, which links Brodmann areas 40 and 45-47 (Rammayya et al. 2010, Rojkova et al. 2016, Barbeau et al 2020). The cortical areas revealed by this VBCM analysis included inferior frontal (BA45,47), and inferior parietal (SMG, BA40) cortices as well as action execution areas involving the primary motor cortex. The cluster identified within the inferior frontal gyrus lesion also overlapped with the uncinate fasciculus linking inferior frontal and temporal areas (Rojkova et al. 2016). This network represents the ‘ventro-dorsal’ subdivision within the dorsal stream, corroborating findings reported in previous lesion mapping studies of apraxia (Pazzaglia et al. 2008, Kalenine et al. 2010, Buxbaum et al. 2014, Hoeren et al. 2014, Dressing et al. 2019), which also parallel areas identified in semantic control in the language literature (Corbett et al. 2009).

On the other hand, the ‘multi-demand sequencing’ factor identified structural correlates involving the left SLF II, which connects the angular gyrus (BA39) with the middle frontal gyrus (BA6)(Rojkova et al. 2016, Barbeau et al. 2020). The significant cluster identified involved the white matter underlying sensorimotor areas and was separable from the previous cluster in that it fell within the ‘dorso-dorsal’ subdivision of the dorsal visual stream (Rizzolatti and Matelli, 2003). Moreover, another cluster was identified with this factor, which included the right (and partly the left) posterior orbitofrontal cortex (pOFC) and underlying anterior cingulum (Rojkova et al. 2016). The pOFC connects to the ventral visual pathway and is described in the literature as being highly multi-modal. This area is involved in sequence learning, decision making and memory retrieval tasks, which are all relevant in naturalistic or multi-step behaviours revealed by this factor (Jackson et al. 2003, Petrides 2007). Furthermore, the pOFC is connected to medial frontal regions for action control. The ventral stream areas it connects to include inferior parietal, parahippocampal and anterior temporal areas (Barbas 2007). The identification of right pOFC was somewhat surprising given the fact our cohort of patients only strokes involving the left hemisphere. The VBCM analysis allows to identify variations in T1 signal intensity that correlate to behaviour. Although this variation is expected to be higher at the lesion site, it is still present in the contra-lesional area. The identification of reduced T1 signal intensity in pOFC within the right hemisphere correlating with deficits in Factor 3 would suggest might play a role in these deficits, possibly through a process known as diaschisis (Marsh et al. 2006, Price et al. 2010).

In conclusion, this study demonstrates the successful implementation of PCA in limb apraxia using a detailed battery of tasks, which elicited three component factors: posture selection, semantic control and multi-demand sequencing. Lesions associated with the latter two components were located within fronto-parietal areas: the ‘ventro-dorsal’ pathway for semantic control and both the dorso-dorsal and anterior cingulum/ventral pathways for multi-demand sequencing.

Limitations: We note that although the sample size for the behaviour analyses is relatively large, only a subset (N=24) had MRI imaging to perform VBCM. Despite identifying significant neural clusters for action semantics and action sequencing, we failed to identify neural correlates for posture selection. Previous studies using similar techniques in apraxia to identify components of motor deficits have either been applied on a small sample of patients (e.g., praxis tasks in Alzheimer’s disease in Gulde et al. 2018) or have compared apraxia with another cognitive deficit such as spatial inattention (Timpert et al. 2015). It is important for future research to replicate and extend the results reported in this study by investigating larger samples of patients. There are two further points that could be addressed in future work. The first should focus on investigating whether the components identified here could be subdivided by including a wider range of tasks such as tasks that involve repetitive movements (e.g., hammering a nail), whether movements requires proximal rather than distal limb control, whether they involve sequential actions (Shallice et al. 1989) and whether they involve communicative gestures. Secondly, it is well known that stroke can result in a wide range of sensory, language and cognitive deficits; therefore, assessing multiple domains simultaneously and then performing a data driven analysis such as PCA would allow us to produce a unified model of stroke deficits and its neural bases. This will help bring these cognitive fields together, even though they have traditionally been studied separately.

## Data Availability

Could be made available anonymised upon request to lead author, depending on research question.

## Acknowledgements

We would like to thank the patients who took part in the study. This work has been supported by a Helen Dawson grant from the British Medical Association, as well as grants from the Oxford University Clinical Academic Graduate School and Oxfordshire Health Services Research Committee to Dr Rounis. Dr Halai and Professor Lambon Ralph were supported by The Rosetrees Trust (no. A1699 to Dr Halai and Professor Lambon Ralph), the ERC (GAP: 670428 – BRAIN2MIND_NEURO_COMP to Professor Lambon Ralph) and MRC intramural funding (MC_UU_00005/18).

## CRediT author contribution statement

**Elisabeth Rounis:** Conceptualisation, Funding Acquisition, Project Administration, Formal Analysis, Writing – Original Draft, Writing – Review & Editing. **Ajay Halai**: Formal analysis, Methodology, Visualization, Validation, Supervision, Analysis tools, Writing - original draft, Writing - review & editing; **Gloria Pizzamiglio**: Project Administration, Data acquisition, Investigation. **Matthew Lambon Ralph**: Conceptualisation, Methodology, Supervision, Writing – Review & Editing.

## Supplemental Material

**Supplementary Table 1.** Individual task scores (normalized out of 100) for all patients in all tasks Those patients who underwent imaging are highlighted in bold (24 in total)

| <u>P</u><br><u>T</u><br><u>N</u><br><u>o</u> | <u>Multi-</u><br><u>Object</u><br><u>use</u> | <u>Go-NoGo</u><br><u>Commissio</u><br><u>n errors at</u><br><u>250ms</u> | <u>Go-</u><br><u>NoGo</u><br><u>Omissio</u><br><u>n errors</u> | <u>Gesture</u><br><u>Recognitio</u><br><u>n</u> | <u>Gesture</u><br><u>Productio</u><br><u>n</u> | <u>Comple</u><br><u>x Figure</u><br><u>Copy</u> | <u>Single</u><br><u>Object</u><br><u>Use</u> | <u>Meaningless</u><br><u>Gesture</u><br><u>Imitation</u> |
| --- | --- | --- | --- | --- | --- | --- | --- | --- |
| **1** | **100** | **80** | **90** | **100** | **100** | **100** | **100** | **90** |
| **2** | **100** | **80** | **80** | **83.33** | **58.33333** | **97.87** | **90** | **70** |
| **3** | **100** | **90** | **76.6666**<br>**7** | **100** | **100** | **89.36** | **100** | **100** |
| 4 | 100 | 100 | 96.6666<br>7 | 100 | 91.66667 | 95.74 | 100 | 0 |
| 5 | 100 | 90 | 90 | 100 | 58.33333 | 97.87 | 90 | 80 |
| **6** | **66.67** | **100** | **90** | **83.33** | **100** | **100** | **90** | **90** |
| 7 | 100 | 60 | 83.3333<br>3 | 100 | 100 | 100 | 100 | 0 |
| 8 | 100 | 100 | 100 | 83.33 | 91.66667 | 85.11 | 83.33 | 100 |
| 9 | 75 | 50 | 50 | 66.67 | 8.333333 | 85.11 | 58.33 | 35 |
| 10 | 100 | 70 | 66.6666<br>7 | 83.33 | 75 | 68.09 | 75 | 75 |
| 11 | 91.67 | 80 | 86.6666<br>7 | 94 | 91.66667 | 89 | 80 | 60 |
| **12** | **100** | **90** | **96.6666**<br>**7** | **100** | **100** | **95.74** | **100** | **100** |
| **13** | **100** | **80** | **93.3333**<br>**3** | **100** | **83.33333** | **87.23** | **100** | **95** |
| **14** | **100** | **100** | **96.6666**<br>**7** | **100** | **91.66667** | **100** | **91.67** | **95** |
| **15** | **100** | **90** | **96.6666**<br>**7** | **100** | **100** | **97.87** | **90** | **100** |
| **16** | **100** | **90** | **90** | **100** | **80** | **93.62** | **100** | **85** |
| **17** | **91.67** | **100** | **100** | **100** | **100** | **100** | **90** | **100** |
| **18** | **100** | **90** | **80** | **100** | **100** | **97.87** | **100** | **95** |
| **19** | **100** | **100** | **100** | **100** | **90** | **97.87** | **100** | **100** |
| 20 | 100 | 100 | 96.6666<br>7 | 83.33 | 80 | 100 | 90 | 85 |
| 21 | 90 | 90 | 96.6666<br>7 | 100 | 75 | 97.87 | 91.67 | 90 |
| 22 | 80 | 80 | 63.3333<br>3 | 100 | 83.33333 | 85.11 | 85 | 80 |
| 23 | 91.67 | 90 | 96.6666<br>7 | 100 | 85 | 100 | 95 | 100 |
| 24 | 100 | 80 | 93.3333<br>3 | 100 | 100 | 97.87 | 100 | 90 |
| 25 | 0 | 100 | 96.6666<br>7 | 100 | 66.66667 | 78.72 | 85 | 80 |
| 26 | 100 | 100 | 93.3333<br>3 | 66.67 | 50 | 85.11 | 85 | 85 |
| 27 | 100 | 89.31818 | 90.5303 | 100 | 100 | 78.72 | 93 | 80 |
| 28 | 80 | 100 | 93.3333<br>3 | 83.33 | 75 | 91.49 | 91.67 | 70 |
| 29 | 83.33 | 100 | 96.6666<br>7 | 100 | 75 | 78.72 | 95 | 100 |
| 30 | 100 | 80 | 86.6666<br>7 | 100 | 95 | 100 | 90 | 85 |
| 31 | 100 | 90 | 93.3333<br>3 | 100 | 100 | 89.36 | 100 | 95 |
| 32 | 91.6 | 70 | 90 | 100 | 91.66667 | 95.74 | 75 | 75 |
| 33 | 100 | 100 | 100 | 100 | 95 | 100 | 91.67 | 80 |
| 34 | 75 | 100 | 100 | 83.33 | 85 | 25.53 | 80 | 70 |
| 35 | 91.67 | 80 | 80 | 100 | 75 | 93.62 | 100 | 70 |
| 36 | 91.67 | 89.31818 | 90.5303 | 83.33 | 90 | 91.49 | 91.67 | 91.67 |
| 37 | 100 | 90 | 90 | 100 | 83.33333 | 78.72 | 90 | 95 |
| 38 | 80 | 80 | 93.3333<br>3 | 83.33 | 90 | 95.74 | 100 | 100 |
| 39 | 100 | 90 | 90 | 100 | 80 | 89.36 | 83.33 | 90 |
| 40 | 100 | 100 | 96.6666<br>7 | 50 | 65 | 91.49 | 91.67 | 90 |
| 41 | 100 | 100 | 100 | 100 | 91.66667 | 93.62 | 85 | 80 |

